# Assessment of knowledge, attitude and practices among Accredited Social Health Activists (ASHAs) towards COVID-19: a descriptive cross-sectional study in Tripura, India

**DOI:** 10.1101/2021.02.17.21251725

**Authors:** Purvita Chowdhury, Subrata Baidya, Debosmita Paul, Pinki Debbarma, Biraj Kalita, Sanjoy Karmakar

## Abstract

With the surge in COVID-19 cases, community healthcare workers (CHW) remain pivotal for proper dissemination of awareness of disease transmission and infection control measures among the communities in low- and middle-income countries. In this context, lack of adequate knowledge and appropriate attitude among the CHW can directly influence the COVID-19 management programme. Therefore, the present study was designed to assess the knowledge, attitude and practices towards COVID-19 among the CHW of India known as Accredited Social Health Activists (ASHAs). A descriptive cross-sectional was conducted in the state of Tripura, Northeast India, among ASHAs with 14-, 4- and 3-item self-administered questionnaire for knowledge, attitude and practice. Around 61.2% of participants had the mean correct answer rate and the mean score of knowledge was 8.57± 2.25 (±SD). As per Bloom’s cut-off, it was observed that only 10% of the ASHAs had adequate knowledge, 30.9% showed positive attitude and 88% adhered to the good practices. It was observed that the indigenous ASHAs were 1.79 times more likely to adhere to the good practice of wearing masks during filed visits in the community (OR: 1.791, 95% CI: 1.059-3.028, p=0.030). Multinomial regression analysis showed that practice was significantly associated with fear of getting infected during service and the community’s fearfulness of ASHAs spreading the disease. Urgent addressing of the provisions of support, guidance and training of grassroot level healthcare workers in rural tough terrains can ensure robust output from the existing community healthcare workers in future pandemic-like emergencies.

## Introduction

The COVID-19 pandemic caused by severe acute coronavirus 2 (SARS-CoV-2) has become one of the most important health problems in recent history affecting more than 50 million people globally (WHO 2020a). After USA, currently India ranks the second highest in COVID-19 caseload (Johns Hopkins University & Medicine 2020). The first case of COVID-19 was detected in India in January when WHO declared the novel Coronavirus outbreak as a Public Health Emergency of International concern.

Current evidences suggest that SARS-CoV-2 is transmitted predominantly from person to person through respiratory secretions in small droplets when in close contact or by touching contaminated surface or objects (WHO 2020b). Pre-existing morbidity, chronic illness and old age have been identified as potential risk factors for disease severity and mortality (Li et al. 2020). Although the public healthcare workers are working tirelessly for COVID-19 management, there is currently no specific antiviral treatment or vaccine available.

The cornerstone of the Indian public healthcare system largely depends on the grassroot level trained female community workers known as Accredited Social Health Activists (ASHAs). In India, ASHAs are the primary frontline workers disseminating awareness and ensuring the recommended preventive measures in the community. They represent an interface between the community and the health facilities forming the primary care delivery system. In this crisis, the WHO recommends preventive measures including social distancing, regular hand washing and maintaining respiratory etiquette (WHO 2020c). With the surge of COVID-19 cases in India, awareness of the disease transmission and infection control measures as implemented by the Govt. of India needs proper dissemination among the communities and this is facilitated by the ASHAs.

It has been observed that most of these community healthcare workers are confronted with a number of problems in course of rendering their duty. The major ones are in the nature of psychological stigma, occupational discrimination and even possible exposure to the disease in the field of duty (WHO 2020d). In addition to this, lack of adequate knowledge has been directly associated with negative attitude, practice and unwillingness to work in the healthcare workers (Aoyagi et al. 2015; McEachan et al. 2016). Therefore, the present study was conducted to assess the knowledge, attitude and practices of ASHAs during COVID-19 pandemic.

## Materials and methods

### Study design

A descriptive cross-sectional study was conducted during July-September 2020 after unlock-1 was declared in India.

### Study setting

During the time of the study, the highest COVID-19 cases were reported from West Tripura District of Tripura. Therefore, four Community healthcare centres under West Tripura district were randomly selected for the study.

### Study participants

The study participants included all the deputed community healthcare volunteers known as Accredited Social Health Activist (ASHA) of the selected CHCs. These ASHAs are a part of National Rural Health Mission, Govt. of India. ASHAs that were unavailable during the period of study due to sickness or were under quarantine were excluded from the study.

### Ethical Clearance

The study has been reviewed and cleared by the Institutional Ethical Committee of the Agartala Government Medical College, Tripura (Ref. No. F (5-234)/AGMC/Academic/IEC Meeting/2020/13149). Participant identity and information confidentiality has been maintained during the study.

### Study tools

A self-administered questionnaire was used for the present study. The questionnaire was developed after reviewing previously conducted studies and consulting the websites of Ministry of Health and Family Welfare (MoHFW), Govt. of India (MoHFW 2020; Zhong et al. 2020).

### Study variables

The questionnaire covered demographic details of the ASHAs like age, education, marital status and ethnicity. Knowledge section of the questionnaire consisted of 14 questions about COVID-19 symptoms, mode of transmission, risk factors and prevention. The answer for each question was either yes, no or don’t know. A correct response was rewarded with 1 point and 0 for an incorrect or ‘don’t know’ response.

The attitude section of the questionnaire comprised of four questions to understand their attitudes towards risk perception and regulations taken by the Government to overcome COVID-19 pandemic situation in the state. This section had responses viz. ‘strongly agree’, ‘agree’, ‘neutral’, ‘don’t agree’ and ‘strongly don’t agree’ weighing a score of 1-5 respectively for correct response.

The practice section comprised of three questions to understand the implementation of the recommended practices in field during the pandemic. This section had responses viz. ‘always, ‘sometimes’ and ‘never’ each weighing 3-1 score for good practice.

### Data Collection

Due to the pandemic situation, an online data collection method using Google online form was developed. The link was distributed to all the Medical Officers along with ASHAs of the selected CHCs and the participants were requested to fill the form. For ASHAs with unavailability of smart phones or internet connectivity issues in rural settings, a printed version of the questionnaire was also distributed to the CHCs. The printed forms were filled up and submitted by the ASHAs during their monthly meeting or ASHA Bharosha Diwas at the CHCs.

### Statistical analysis

Data was coded and analysed using SPSS v25(IBM Corp., Armonk, NY, USA) and GraphPad Prism5. Descriptive data was summarised as Mean ± standard deviation and categorical variables were summarised as proportions and frequencies. Sufficient knowledge, positive attitude and good practice were considered according to Bloom’s cut-off of 80% (Kaliyaperumal 2004). Logistic regression and multinomial logistic regression were used to determine the factors associated with knowledge, attitude and practice with COVID-19. Statistical significance of each of the predictor variables on the outcome variables was presented by their odds ratio (OR) values with a 95% confidence interval (95% CI) at p<0.05.

## Results

Of the 240 ASHAs approached, a total of 210 completed the study with a response rate of 87.5%. The socio-demographic features of the study participants are shown in table 1. The mean age of the participating ASHAs was 39.95±7.27 (±SD) years where more than half of the participants belonged to the age group of 36-45 years (n=109, 58.9%). Around 72% (n=151) of the ASHAs had high school or higher level of formal education and 5.2% (n=11) had less than primary level education. The study group comprised 40.5% (n=85) of indigenous population and majority of the participants were married (n=190, 90%).

**Table 1:**
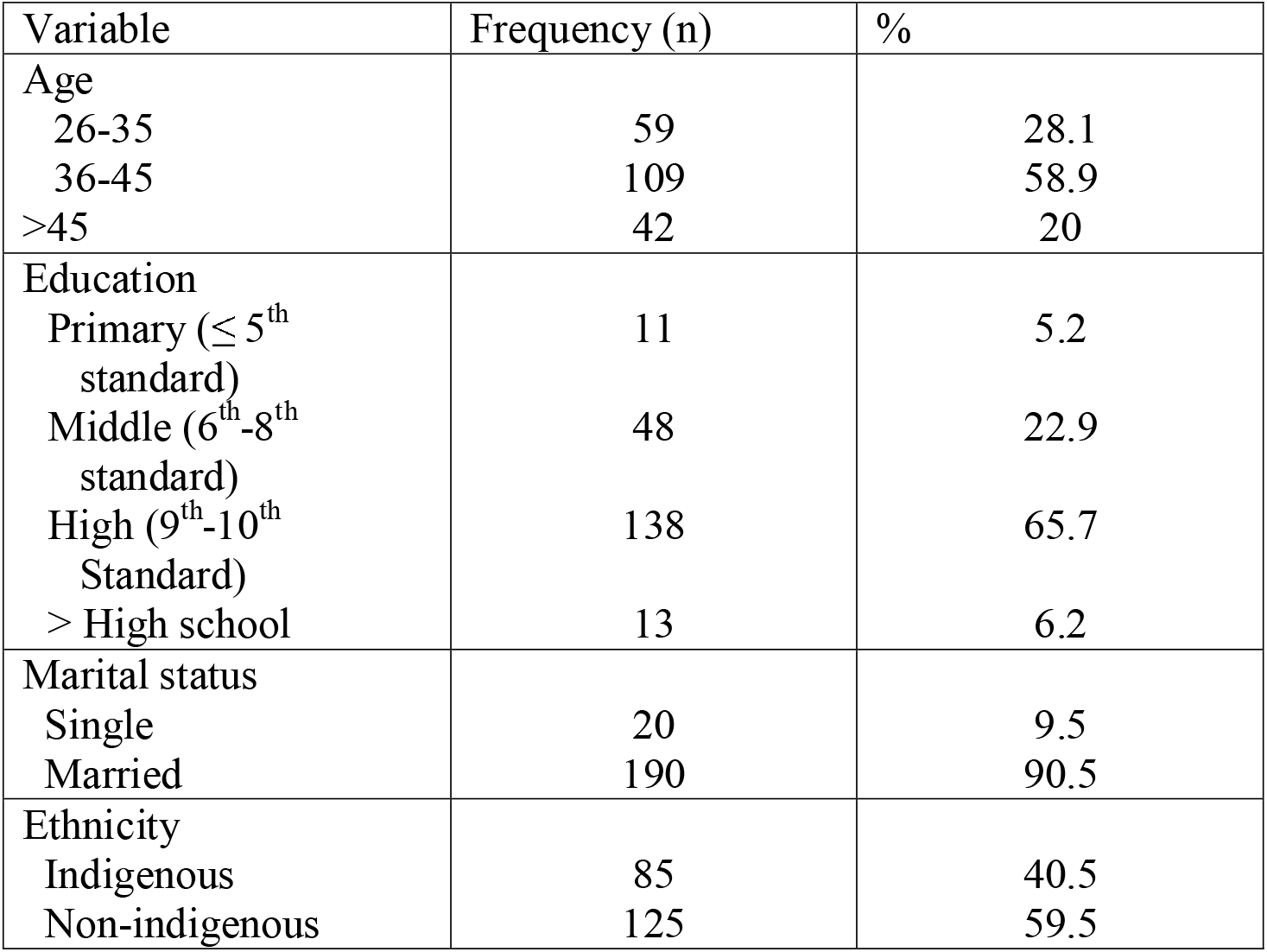
Socio-demographic features of studied participants (N=210)

| Variable | Frequency (n) | % |
| --- | --- | --- |
| Age |  |  |
| 26-35 | 59 | 28.1 |
| 36-45 | 109 | 58.9 |
| >45 | 42 | 20 |
| Education |  |  |
| Primary ($\leq 5^{\text{th}}$ standard) | 11 | 5.2 |
| Middle ($6^{\text{th}}$-$8^{\text{th}}$ standard) | 48 | 22.9 |
| High ($9^{\text{th}}$-$10^{\text{th}}$ Standard) | 138 | 65.7 |
| > High school | 13 | 6.2 |
| Marital status |  |  |
| Single | 20 | 9.5 |
| Married | 190 | 90.5 |
| Ethnicity |  |  |
| Indigenous | 85 | 40.5 |
| Non-indigenous | 125 | 59.5 |

### Knowledge

Knowledge among the ASHA workers about COVID-19 symptoms, transmission and preventive measures are described in Table 2. The mean score of knowledge among the participants was 8.57± 2.25 (±SD) with a score range of 2 to 13. Around 61.22% of participants had the mean correct answer rate. Majority (90%) of the participants were aware of the primary symptoms for COVID-19. However, only one third of the total participants had the perception that in summers, high temperatures could not kill the virus.

**Table 2:**
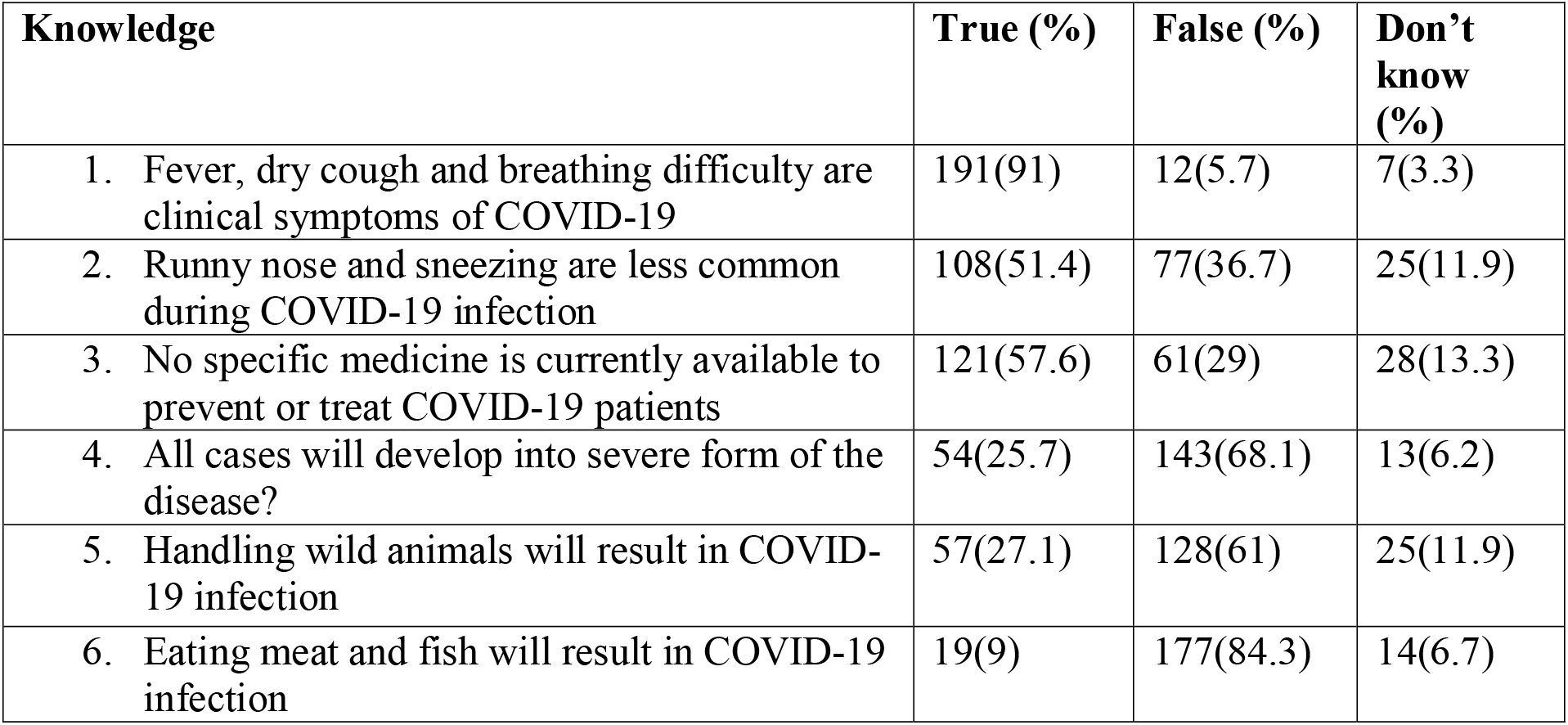
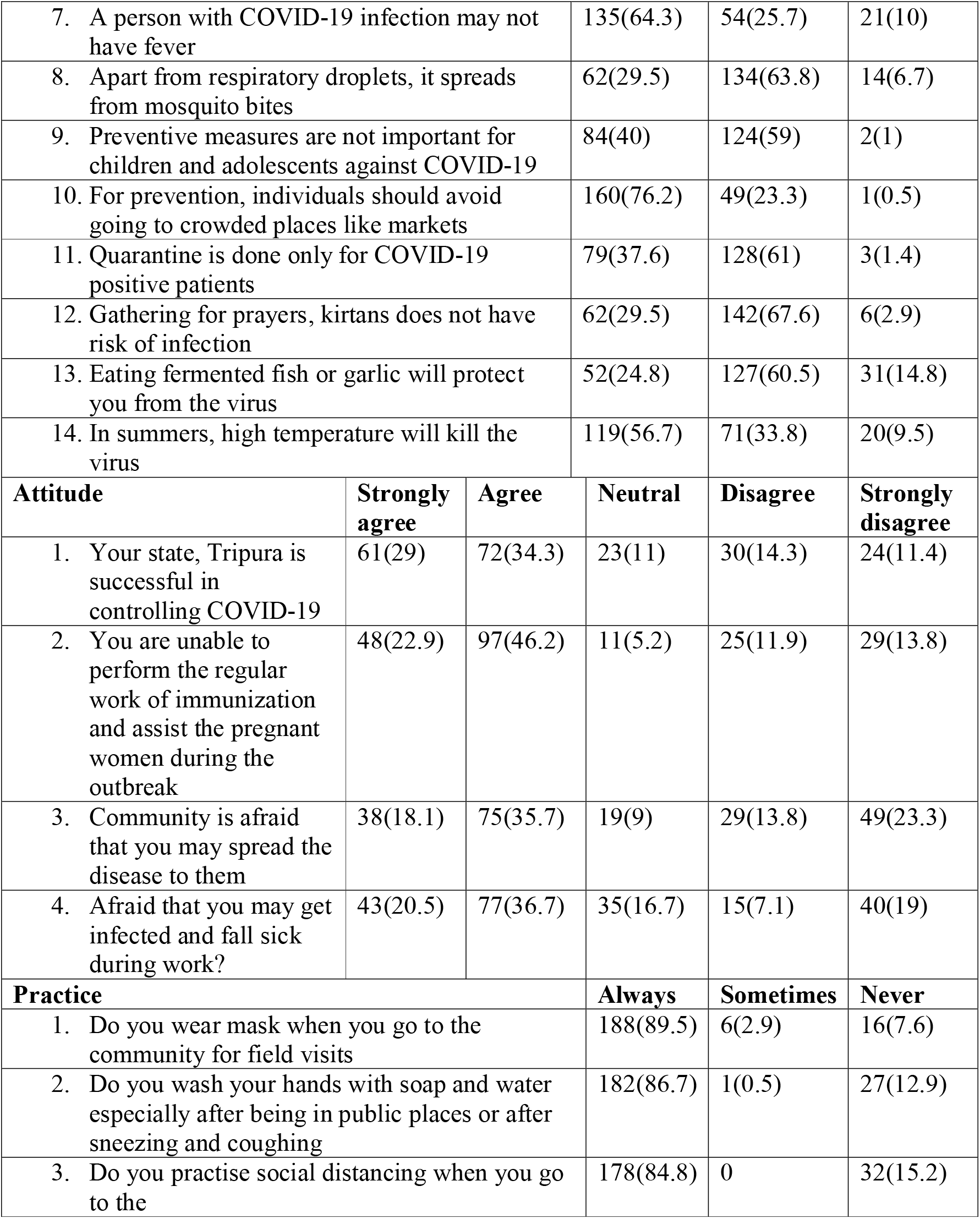
Knowledge attitude and practices (KAP) towards COVID-19 among the ASHAs of Tripura

Unfortunately, 40% (n=84) of the ASHAs thought that prevention measures for children and adolescent are not essential. About 57% (n=121) correctly acknowledged that there was currently no specific medicine for the disease while 13.3% (n=28) were uncertain. In fact, only 10.4% (n=22) of the ASHAs had sufficient/good knowledge (knowledge score ≥80%) regarding COVID-19 and the highest proportion of good knowledge was observed among the ASHAs of 36-45(68.2%) age group (Table 3).

**Table 3:**
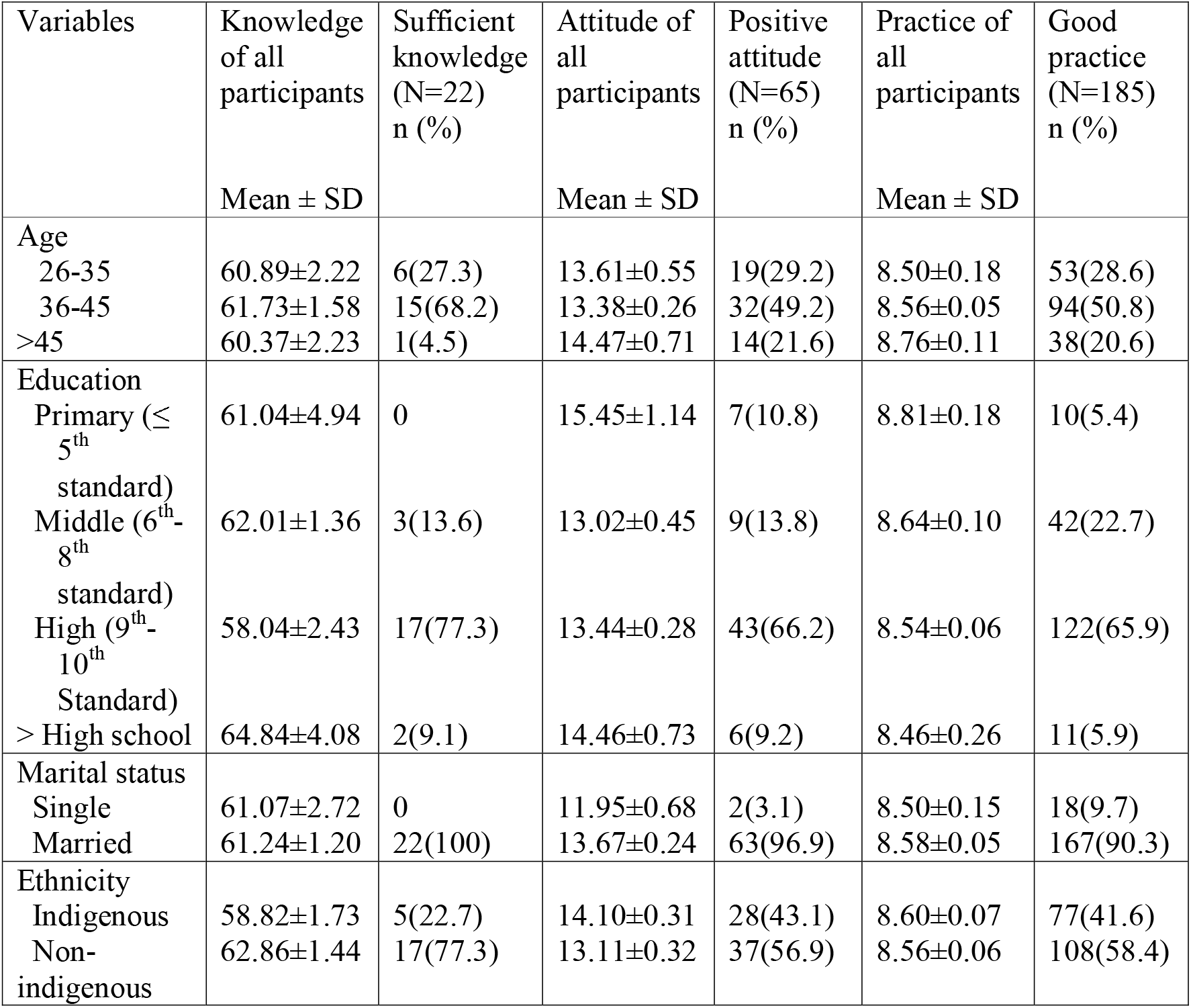
KAP towards COVID-19 of the ASHAs according to their socio-demographic features

| Variables | Knowledge of all participants<br>Mean ± SD | Sufficient knowledge (N=22)<br>n (%) | Attitude of all participants<br>Mean ± SD | Positive attitude (N=65)<br>n (%) | Practice of all participants<br>Mean ± SD | Good practice (N=185)<br>n (%) |
| --- | --- | --- | --- | --- | --- | --- |
| Age |  |  |  |  |  |  |
| 26-35 | 60.89±2.22 | 6(27.3) | 13.61±0.55 | 19(29.2) | 8.50±0.18 | 53(28.6) |
| 36-45 | 61.73±1.58 | 15(68.2) | 13.38±0.26 | 32(49.2) | 8.56±0.05 | 94(50.8) |
| >45 | 60.37±2.23 | 1(4.5) | 14.47±0.71 | 14(21.6) | 8.76±0.11 | 38(20.6) |
| Education |  |  |  |  |  |  |
| Primary (≤ 5<sup>th</sup> standard) | 61.04±4.94 | 0 | 15.45±1.14 | 7(10.8) | 8.81±0.18 | 10(5.4) |
| Middle (6<sup>th</sup>-8<sup>th</sup> standard) | 62.01±1.36 | 3(13.6) | 13.02±0.45 | 9(13.8) | 8.64±0.10 | 42(22.7) |
| High (9<sup>th</sup>-10<sup>th</sup> Standard) | 58.04±2.43 | 17(77.3) | 13.44±0.28 | 43(66.2) | 8.54±0.06 | 122(65.9) |
| > High school | 64.84±4.08 | 2(9.1) | 14.46±0.73 | 6(9.2) | 8.46±0.26 | 11(5.9) |
| Marital status |  |  |  |  |  |  |
| Single | 61.07±2.72 | 0 | 11.95±0.68 | 2(3.1) | 8.50±0.15 | 18(9.7) |
| Married | 61.24±1.20 | 22(100) | 13.67±0.24 | 63(96.9) | 8.58±0.05 | 167(90.3) |
| Ethnicity |  |  |  |  |  |  |
| Indigenous | 58.82±1.73 | 5(22.7) | 14.10±0.31 | 28(43.1) | 8.60±0.07 | 77(41.6) |
| Non-indigenous | 62.86±1.44 | 17(77.3) | 13.11±0.32 | 37(56.9) | 8.56±0.06 | 108(58.4) |

Although the mean knowledge score of ASHAs with education higher than high school was more than the rest of the participants, no significant association was observed between them as well as all the other independent variables (Figure 1).

**Fig. 1.**
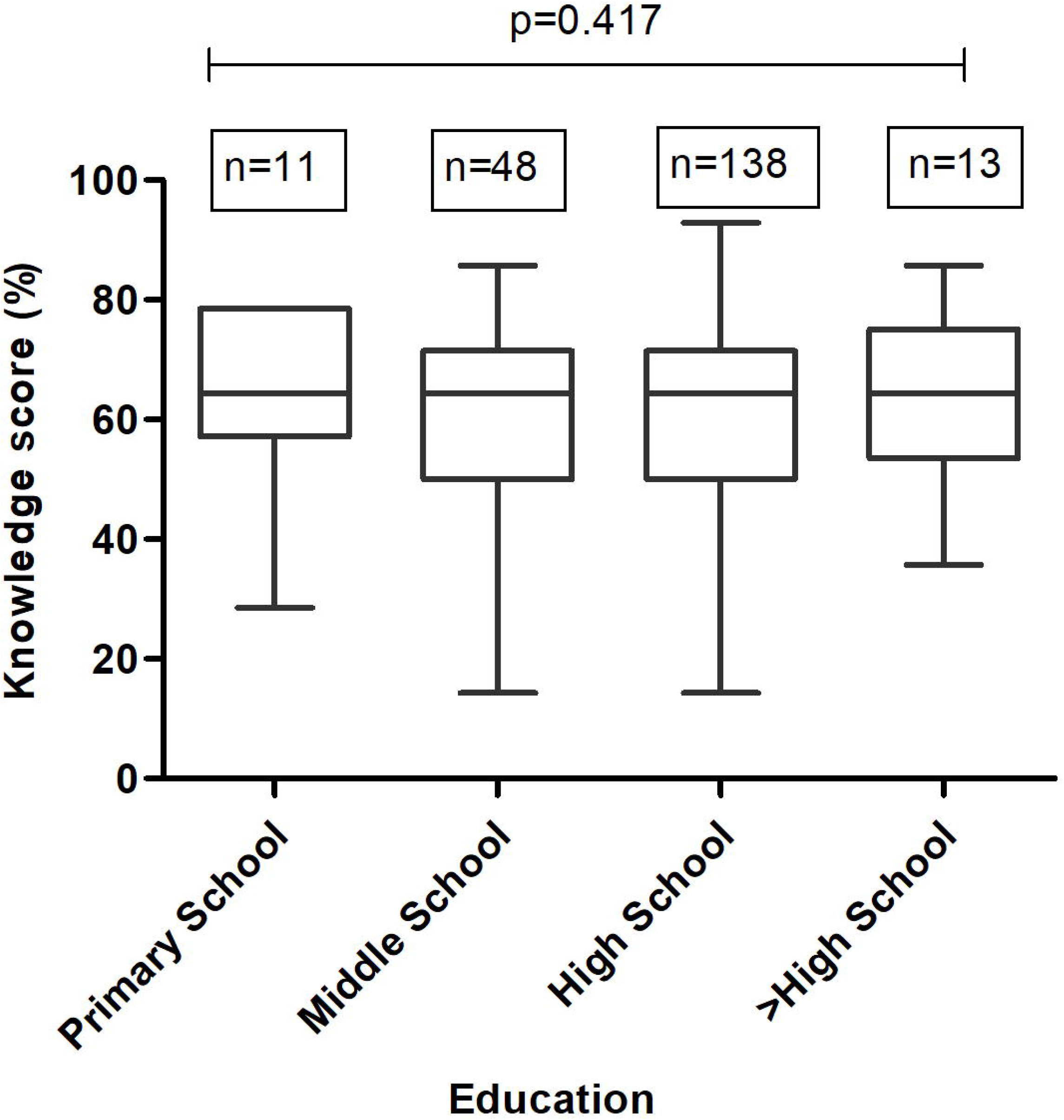
Knowledge score among the studied participants stratified according to their education

### Attitude

The mean attitude score was found to be 13.51 ± 3.34 (± SD) and positive attitude was observed among 30.9% (n=65) of the ASHAs. As seen in table 2, nearly 63% (n=133) of the participants believed that Tripura was successful in controlling the pandemic situation in the state. Table 3 shows the mean attitude score was seen highest among 49.2% ASHAs of 36-45 group. Around 69% (n=145) of the ASHAs agreed that the pandemic situation caused disruption in providing routine immunization for children and health services to pregnant women. The mean attitude score was significantly higher among the married than the single ASHAs (p=0.04) (Table 4).

**Table 4:**
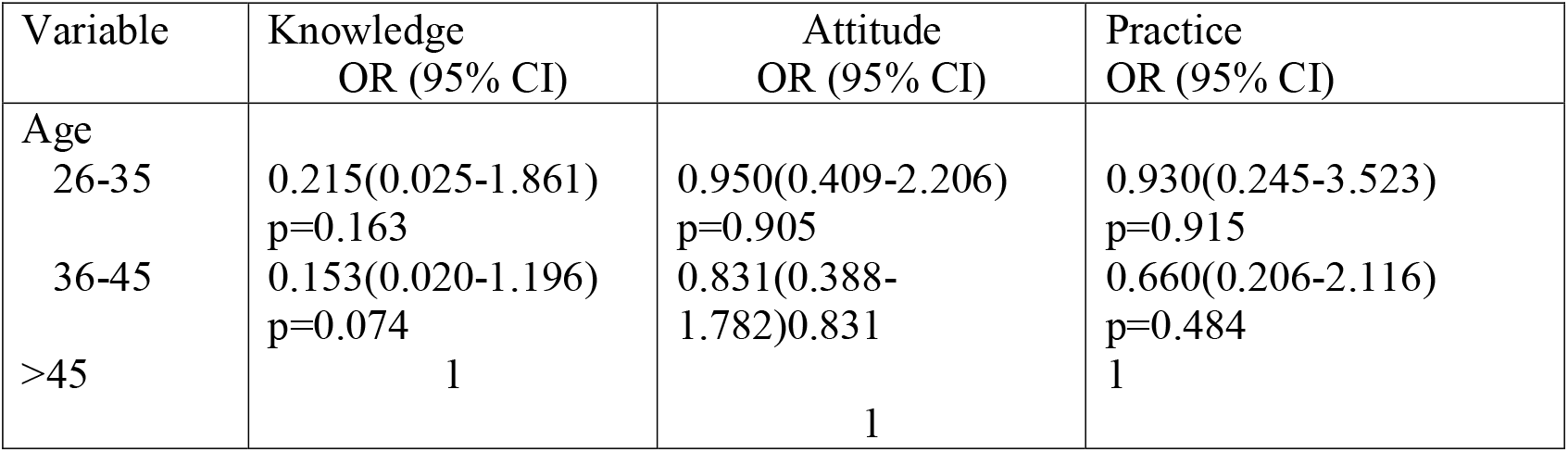
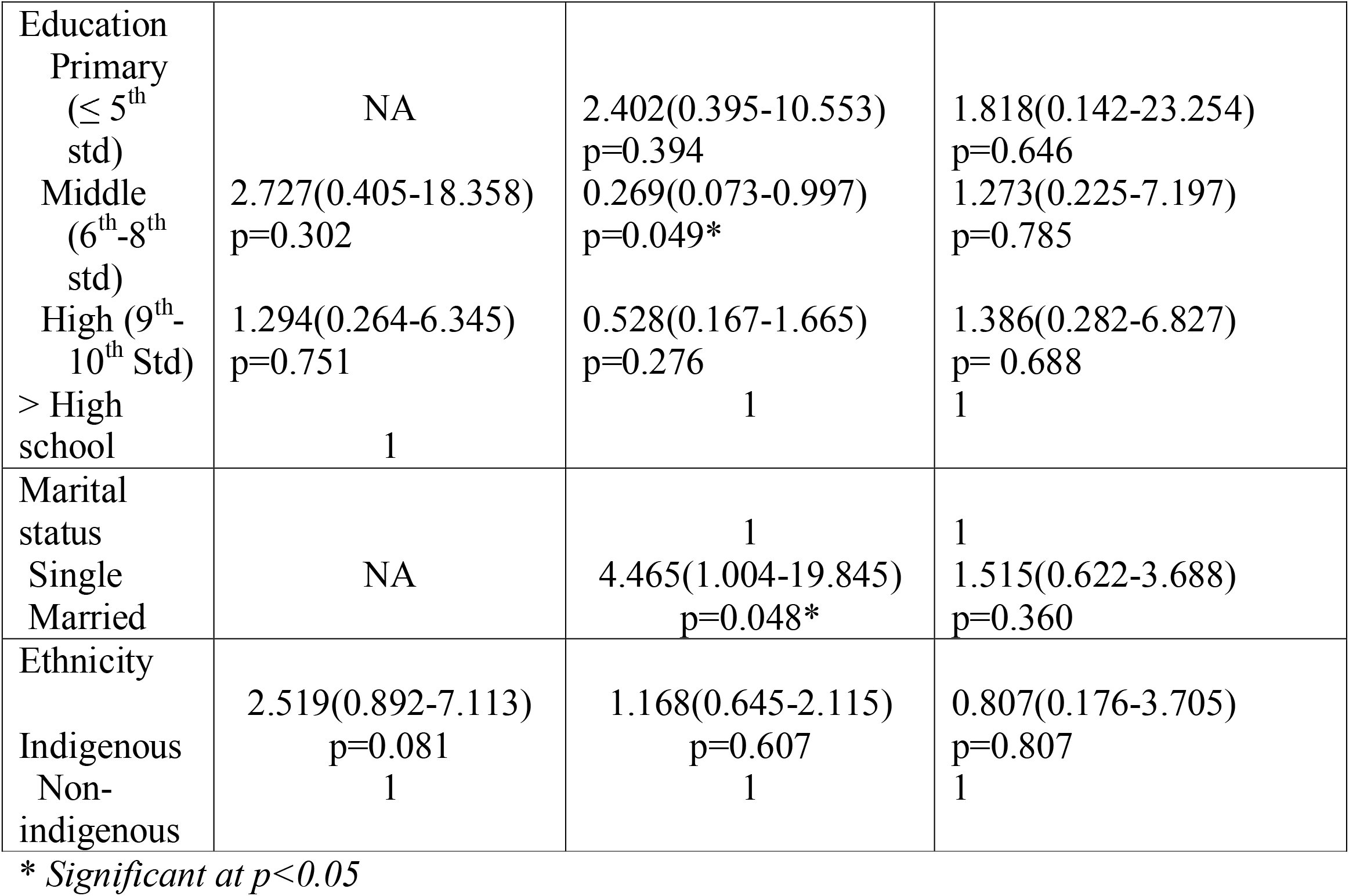
Factors associated with KAP among the ASHAs of Tripura

The fear of being infected with the virus while working in the field was iterated by 57.1% (n=120) ASHAs. When asked about community’s perception about them, 53.8% agreed that community was apprehensive that ASHAs would spread COVID-19 during their field visits. The age groups of <45 years are less likely to have good attitude in believing that their state is successful in controlling the disease (Table 5).

**Table 5:** Factors for attitude and practice towards COVID-19 among the ASHAs

|  | **Attitude** |  |  |  | **Practice** |  |  |
| --- | --- | --- | --- | --- | --- | --- | --- |
|  | Your state, Tripura is successful in controlling COVID-19 | You are unable to perform the regular work of immunization and assist the pregnant women during the outbreak | Community is afraid that you may spread the disease to them | Afraid that you may get infected and fall sick during work? | Do you wear mask when you go to the community for field visits | Do you wash your hands with soap and water especially after being in public places or after sneezing and coughing | Do you practise social distancing when you go to the |
| **Age** |  |  |  |  |  |  |  |
| 26-35 | 0.172(0.040-0.732)<br>p=0.017 | 1.789(0.254-12.578)<br>p=0.559 | 0.979(0.213-4.513)<br>p=0.978 | 1.325(0.435-4.031)<br>p=0.620 | 1.815(0.934-3.529)<br>p=0.07 | 0.588(0.326-1.060)<br>p=0.588 | 1.502(0.922-2.445)<br>p=0.105 |
| 36-45 | 0.352(0.124-1.000)<br>p=0.05* | 1.616(0.290-9.010)<br>p=0.584 | 1.356(0.381-4.826)<br>p=0.638 | 1.173(0.432-3.181)<br>p=0.754 | 1.660(0.831-3.198)<br>p=0.130 | 0.901(0.548-1.480)<br>p=0.680 | 1.127(0.680-1.867)<br>p=0.642 |
| >45 | 1 | 1 | 1 | 1 | 1 | 1 | 1 |
| **Education** |  |  |  |  |  |  |  |
| Primary (≤ 5<sup>th</sup> standard) | NA | 3.335(0.273-40.802)<br>p=0.346 | 2.065(0.150-28.448)<br>p=0.588 | 0.474(0.066-3.401)<br>=0.458 | 1 | 1 | 1 |
| Middle (6<sup>th</sup>- | NA | 0.101(0.005-2.136) | 1.167(0.117- | 0.310(0.072- | 0.681(0.184- | 0.657(0.190-2.269) | NA |
| 8<sup>th</sup> standard) |  | p=0.141 | 11.581)<br>p=0.895 | 1.336)<br>p=0.116 | 2.523)<br>p=0.565 | p=0.567 |  |
| High (9<sup>th</sup>-10<sup>th</sup> Standard) | NA | 0.325(0.032-3.298)<br>p=0.342 | 0.756(0.085-6.710)<br>p=0.802 | 0.341(0.094-1.231)<br>p=0.100 | 1.037(0.330-3.257)<br>p=0.957 | 1.038(0.346-3.113)<br>p=0.943 | NA |
| > High school |  | 1 | 1 | 1 | NA | NA | NA |
| **Marital status** |  |  |  |  |  |  |  |
| Single | 1 | 1 | 1 | 1 | 1 | 1 | 1 |
| Married | 0.765(0.187-3.133)<br>p=0.710 | 1.217(0.140-10.456)<br>p=0.859 | 0.560(0.141-2.226)<br>p=0.460 | 0.853(0.258-2.819)<br>p=0.794 | 1.612(0.538-4.831)<br>p=0.394 | 0.617(0.338-1.127)<br>p=0.116 | 0.952(0.504-1.798)<br>p=0.879 |
| **Ethnicity** |  |  |  |  |  |  |  |
| Indigenous | 1.084(0.415-2.826)<br>p=0.870 | 0.549(0.134-2.243)<br>p=0.403 | 0.334(0.102-1.087)<br>p=0.06 | 1.138(0.863-3.911)<br>p=0.114 | 1.791(1.059-3.028)<br>p=0.030* | 0.734(0.457-1.177)<br>p=0.199 | 0.805(0.524-1.235)<br>p=0.320 |
| Non-indigenous | 1 | 1 | 1 | 1 | 1 | 1 | 1 |

*\*Significant at $p < 0.05$*

### Practice

More than 85% (n=185) of the ASHAs had good practices. Up to 89% of the ASHAs affirmed that they always wear masks when visiting the field. Of the surveyed participants, 86.7% ASHAs reportedly always washed their hands with soap and water after coming back from public place. No significant association was observed between the independent variables and the questions of practice under the study. However, multinomial logistic regression showed that attitude was significantly associated with the practices of the ASHAs. From table 5, it was observed that the indigenous ASHAs were 1.79 times more likely to adhere to the good practice of wearing masks during filed visits in the community (OR: 1.791, 95% CI: 1.059-3.028, p=0.030). Multinomial regression analysis showed that practice was significantly associated with fear of getting infected during service and the community’s fearfulness of ASHAs spreading the disease (Table 6).

**Table 6:** Fearfulness towards COVID-19 among the ASHAs

|  | Community is afraid that you may spread the disease to them | Afraid that you may get infected and fall sick during work? |
| --- | --- | --- |
| Knowledge Score [Good($\geq 80\%$ bloom cutoff) vs poor($< 80\%$)] | 0.963(0.705-1.315)<br>p=0.811 | 1.012(0.730-1.405)<br>p=0.942 |
| Practice Score [Good($\geq 80\%$ bloom cutoff) vs poor($< 80\%$)] | 2.244(1.641-3.067)<br>p=0.001* | 2.461(1.710-3.542)<br>p=0.001* |

\*Significant at $p < 0.05$

## Discussion

With over 9,00,000 engaged ASHAs (January 2019), the community health workers (CHW) known as ASHAs in India have come a long way since its initiation in 2005, forming a sustainable frontline workforce of the Indian public healthcare system (National Health Systems Resource Centre 2019). In fact, in response to the COVID-19 pandemic, prioritizing in CHW has been suggested as one of the most equitable and efficient ways to uphold the stumbling healthcare system around the world (Bose 2020; Peretz et al. 2020). Therefore, critical assessment of adequate knowledge on transmission, clinical presentations, prevention strategies among the CHW including ASHAs of India are indispensable during the pandemic. From the present study, it was observed that only 10% of the ASHAs had adequate knowledge, 30.9% showed positive attitude and 88% adhered to the good practices.

To the best of our knowledge, this is the first study to assess the KAP among ASHAs with respect to COVID-19 pandemic in India. The mean score of knowledge as observed from the study was comparatively lower than that reported among public of Saudi Arabia (Al-Hanawi et al. 2020). Overall, 91% of the ASHAs were aware of the symptoms which corroborates with a study in Pakistan where 84% of the primary healthcare workers were aware of the signs and symptoms of COVID-19 (Saqlain et al. 2020). Correct rate of response of 61.2% was in line with a study conducted in Nepal (range: 30-99%) (Paudel et al. 2020). In contrast previous studies among Indian adults with type 1 diabetes mellitus and Indian residents reported 85% and 74% for overall correct rate (Kartheek et al. 2020; Pal et al. 2020).

In the present study, a relatively small (10%) proportion of the ASHAs had adequate knowledge about COVID-19. This is dramatically lower than the adequate knowledge among healthcare workers from Uganda (69%) and Nepal (83.1%) (Olum et al. 2020; Paudel et al. 2020). Another recent study conducted in Mumbai, India reported 71.2% adequate knowledge among healthcare professionals (Modi et al. 2020). This is probably because the other studies were conducted among a wide range of healthcare professionals including doctors, pharmacists and nurses who are highly educated and have technical proficiencies. On the other hand, our study involves community women volunteers who require a minimum qualification of 10^th^ standard to be engaged as ASHAs. The literacy of the ASHAs in different areas may also vary and the education criterion is also relaxed in areas where a suitable woman is not available.

Overall, more than 60% of the ASHAs had confidence that their state is successful in overcoming the pandemic situation which is consistent with a previous finding among Egyptian doctors (40%) (Abdel Wahed et al. 2020). The optimistic belief of the ASHAs might have been enhanced by the prompt COVID-19 pandemic response by the Indian Government ensuring the world’s largest lockdown, and relatively lower number of cases in Tripura than rest of India during the study period (Coronavirus outbreak in India 2020; Venkata-Subramani and Roman 2020). Our findings have shown that marital status has been significant in affecting a positive attitude in the ASHAs. The attitude of healthcare workers is the result of various psychological dimensions. In fact, studies have claimed that highest levels of “burnout” among healthcare workers have been significantly associated with being single or divorced which has direct consequences on the care quality (Cañadas-De la Fuente et al. 2018). Although no significant association was observed between knowledge score and attitude during the study, the importance of adequate knowledge on prevention and coping strategies cannot be ignored (McEachan et al. 2016). Understanding the risk perception among CHW is crucial for improving the attitude and ensuring healthy workplace (CDC 2020). More than 50% of the ASHAs feared of getting infected and being implicated as disease spreaders in the community. In contrast, previous studies report that 92%, 83.8% and 85% of the HCW with high knowledge scores feared being infected in the field (Kandasamy et al. 2020; Maleki et al. 2020; Zhang et al. 2020).

During the pandemic, around 90% of the affected countries have reported disruptions in routine healthcare services (immunization, maternal antenatal check-ups) which were also reiterated by the study participants (WHO 2020e). This situation is a learning lesson for the healthcare system and warrants improved preparedness for emergencies.

Irrespective of knowledge scores and demographic characteristics, more than 80% of ASHAs affirmed of practicing appropriate habits like wearing masks, washing hands and maintaining social distance during field visits. This finding aligns with a previous study among Chinese healthcare workers which reports 89.7% good practices towards COVID-19 (Zhang et al. 2020). Importantly, the adherence to good practices is affected by the attitude of healthcare workers which is observed in the present study and is comparable to a study conducted in Nepal (Limbu et al. 2020).

## Limitations

There are a few limitations in the study. The study was conducted after the lockdown was lifted in India and so the responses of the ASHAs may be biased in respect to their practices in the field during lockdown. Secondly, the study was conducted in one of the highest covid-19 reporting districts of Tripura which may not reflect the KAP of entire country. Nevertheless, to the best of our knowledge, this is the first study to determine KAP towards COVID-19 among the ASHAs of India.

## Conclusion

Although technical guidelines have been issued by the Government of India, dissemination of this information and proper training are crucial during emergencies like the present pandemic. Future research is urgently needed in addressing the provisions of support, guidance and training of grassroot level healthcare workers in rural tough terrains with poor phone and internet connectivity; it still remains a tangible challenge in pandemic scenario. We suggest that a provision of Government recognition and extending support in countering psychological stigma and ostracization faced by ASHAs can go a long way in ensuring robust output from the existing community healthcare workers in future pandemic-like emergencies.

## Data Availability

All the data generated during the study are available in the supplementary file submitted with the manuscript.

## Declarations

### Ethical Approval

The study was performed in line with the principles of the Declaration of Helsinki and approved by the Institutional Ethics Committee of Agartala Government Medical College, Tripura, India.

### Consent to participate

Informed consent was obtained from all individual participants included in the study.

### Consent for publication

Not applicable

### Availability of data and material

All the data generated during the study are available in the *Supplementary File* submitted with the manuscript.

### Code availability

Not applicable

### Authors’ contributions

The study was designed and conceived by Subrata Baidya and Purvita Chowdhury. Data collection was facilitated by Purvita Chowdhury, Debosmita Paul, Pinki Debbarma and Sanjoy Karmakar. Data was analysed by Biraj Kalita. The manuscript was written and edited by Purvita Chowdhury and Subrata Baidya. All the authors approve the final version of the manuscript and agree to be accountable on all aspects of the manuscript.

